# Artificial Intelligence in Medicine: Revolutionizing Healthcare Practices and Patient Outcomes

**DOI:** 10.1101/2025.03.23.25324467

**Authors:** Yudi Kurniawan Budi Susilo, Shamima Abdul Rahman, Dewi Yuliana, Faradiba Abdul Rasyid

**Affiliations:** Faculty of Business and Technology, University of Cyberjaya, 63000 Cyberjaya Selangor, Malaysia; Graduate Research School, University of Cyberjaya, 63000 Cyberjaya Selangor, Malaysia; Faculty of Pharmacy, Universitas Muslim Indonesia, Makassar, Indonesia

**Keywords:** Artificial Intelligence, Personalized Medicine, Healthcare Innovation, AI-Driven Diagnostics, Machine Learning, Deep Learning, Predictive Analytics

## Abstract

Artificial intelligence (AI) has transformed medicine, advancing diagnostics, treatment, and patient outcomes. This study employs bibliometric analysis and the PRISMA framework to explore AI research trends in medicine from 2019 to 2023. Key findings reveal significant growth in publications, with radiomics, genomics, and predictive analytics as major focus areas. Federated learning frameworks and wearable technologies emerge as critical innovations, addressing challenges such as data privacy and real-time monitoring. Despite these advancements, significant barriers persist, including algorithmic bias, data confidentiality, and the need for infrastructure and training to integrate AI into clinical workflows. Future directions emphasize the importance of interdisciplinary collaboration, ethical AI development, and explainable models to ensure trust and equitable access. By addressing these challenges, AI has the potential to revolutionize healthcare systems, optimize resource allocation, and enhance personalized medicine. This study highlights the transformative role of AI in shaping the future of medicine and improving global health outcomes.

## 1.0 INTRODUCTION

Artificial intelligence (AI) has rapidly emerged as a transformative force in medicine, fundamentally reshaping healthcare practices and radically enhancing patient outcomes. The integration of advanced AI algorithms into clinical settings has spurred significant improvements in diagnostic accuracy, streamlined operational workflows, and personalized treatment approaches, demonstrating the potential to revolutionize the way healthcare is delivered [1], [2]. In radiology particularly, AI-driven image analysis and decision support systems have demonstrated greater precision in detecting abnormalities, thereby accelerating early disease detection and intervention [1], [3]. This evolution from traditional practices to data-driven methodologies underscores the significance of AI as not merely a tool but also a catalyst for change within medical practice [4], [5]. As such, the interplay between computational intelligence and clinical expertise forms the cornerstone of this transformative era in medicine [2], [6].

## 2.0 LITERATURE

The historical evolution of AI in healthcare has witnessed a leap from rudimentary rule-based systems to sophisticated deep learning frameworks that emulate complex human cognitive functions. Researchers have highlighted that early AI applications contributed modestly to clinical decision-making; however, recent innovations have drastically expanded the frontiers of diagnostic and therapeutic paradigms [4], [7]. Machine learning algorithms, empowered by massive datasets and robust computing power, are now capable of uncovering hidden patterns within clinical data, leading to more accurate predictions and individualized care strategies [3], [8]. These technological advancements, as documented in various reviews and scientometric analyses, demonstrate the transformative promise of AI in improving patient outcomes across multiple medical disciplines [6], [9]. Consequently, AI has evolved from a theoretical concept to a practical, indispensable asset in contemporary healthcare practice [10], [11].

The application domain of AI in medicine spans an extensive range of specialties, including radiology, ophthalmology, anesthesiology, and even nursing, each being reshaped by its unique innovations. In radiology, AI has been influential in optimizing workflow efficiency while enhancing diagnostic capabilities through automation and deep data analyses [1], [12]. Similarly, in ophthalmology, AI-powered imaging systems have improved the early detection of ocular diseases, thereby facilitating timely treatment interventions and reducing vision loss risks [13], [8]. Complementarily, in anesthesiology, predictive models have been used to foresee patient responses to anesthetic agents, marking an evolution towards more personalized and safe perioperative care [14], [15]. Such cross-disciplinary applications illustrate the multifaceted role of AI in enhancing healthcare delivery and elevating clinical practice standards [2], [16].

Alongside diagnostic and surgical improvements, AI has also made significant inroads in clinical decision-making processes. Machine learning techniques have empowered clinicians to analyze vast amounts of unstructured and structured data, enabling them to make evidence-based decisions regarding patient management [4], [3]. AI-driven risk-prediction models have become central in identifying high-risk patient populations, thereby allowing for preemptive measures and tailored clinical interventions [4], [8]. Moreover, these predictive analytics have been integrated into perioperative settings to optimize treatment plans and mitigate potential complications [17], [15]. The analytical prowess of AI thus serves as a powerful adjunct to the clinical acumen of healthcare professionals, enhancing the overall quality of patient care [5], [9].

Contemporary literature repeatedly emphasizes the transformative potential of AI in streamlining healthcare workflows and supporting clinical decision-making. In various medical fields, including oncology, cardiovascular medicine, and neurology, AI systems analyze complex datasets to uncover diagnostic insights often missed by human evaluation alone [18], [5]. Deep learning models have demonstrated particularly high efficacy in processing multimodal data—ranging from imaging to genomic sequences—thereby bridging the gap between biological complexity and clinical interpretation [8], [7]. Furthermore, the integration of AI in routine clinical practice has paved the way for real-time decision support systems, amplifying the capacity of clinicians to deliver personalized care [17], [19]. Researchers contend that this continuous interplay between AI-driven analysis and clinician expertise is imperative for realizing the full potential of precision medicine [4], [8].

Ethical considerations and regulatory challenges form a critical aspect of the discourse on AI in medicine, as its rapid adoption necessitates rigorous oversight. The ethical dimensions of algorithmic bias, data privacy, and transparency in AI systems have been extensively discussed, highlighting the importance of responsible and equitable AI deployment [20], [21]. Scholars advocate for collaborative frameworks involving healthcare providers, patient advocacy groups, and regulatory bodies to ensure that AI technologies are validated rigorously while upholding the highest standards of patient safety [1], [20]. There are also calls for constant monitoring and evaluation of AI systems to prevent inadvertent harm due to errors in algorithmic predictions, which further emphasizes the need for strong governance structures [21], [22]. These ethical and regulatory imperatives underpin the responsible integration of AI into clinical practice, ensuring that the technology augments rather than undermines patient trust [17], [23].

## 3.0 METHODS

This study employs a bibliometric analysis to explore the evolution, trends, and contributions of research on artificial intelligence (AI) in medicine, focusing on its impact on healthcare practices and patient outcomes. The Preferred Reporting Items for Systematic Reviews and Meta-Analyses (PRISMA) framework was used to ensure a rigorous and systematic approach to data collection, screening, and analysis.

**Figure 1.**
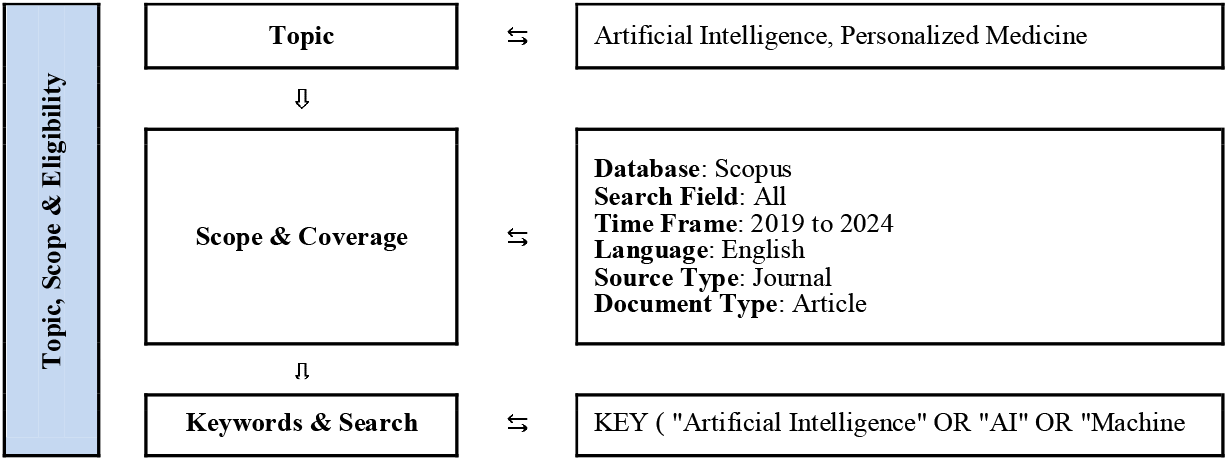
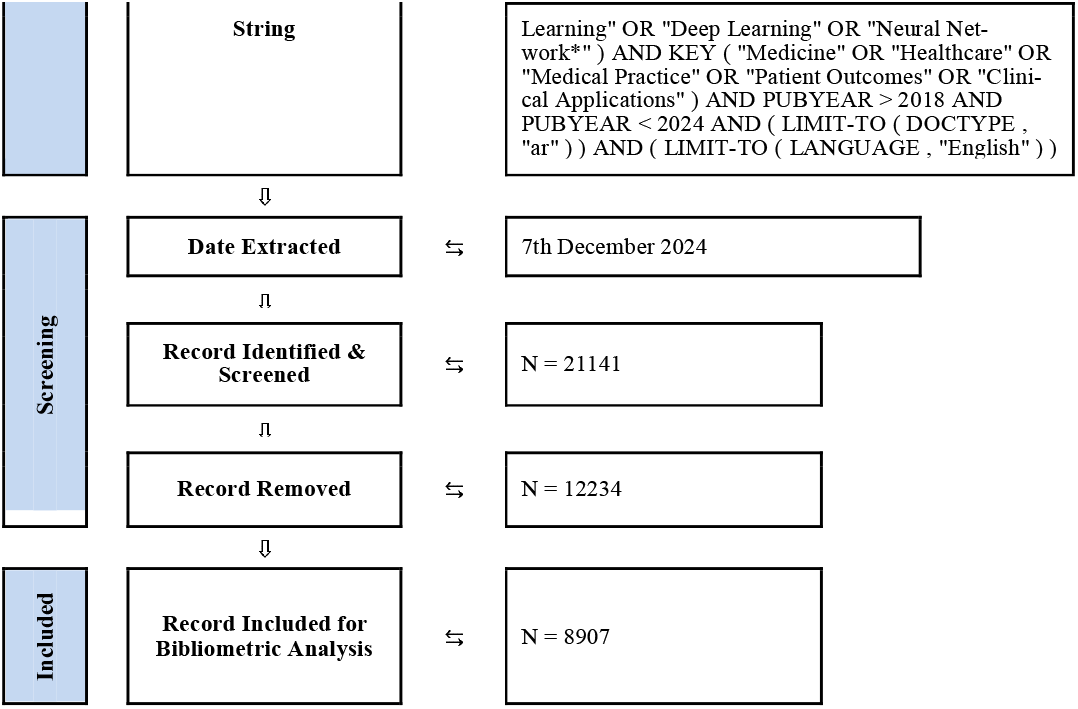
Flow diagram of the search strategy in exploring the evolution, trends, and contributions of research on artificial intelligence (AI) in medicine

The research topic, “Artificial Intelligence in Medicine: Revolutionizing Healthcare Practices and Patient Outcomes,” was defined to explore the interdisciplinary applications of AI in healthcare. The scope was designed to cover global publications indexed in the Scopus database, ensuring comprehensive coverage of the field. The study considered research published between 2019 and 2024, limiting the dataset to journal articles in the English language.

A detailed search string was developed to identify relevant studies, incorporating keywords such as “Artificial Intelligence,” “AI,” “Machine Learning,” “Deep Learning,” “Neural Network,” “Medicine,” and “Healthcare.” Boolean operators and field-specific filters were used to enhance precision and focus:

KEY (“Artificial Intelligence” OR “AI” OR “Machine Learning” OR “Deep Learning” OR “Neural Network*”) AND KEY (“Medicine” OR “Healthcare” OR “Medical Practice” OR “Patient Outcomes” OR “Clinical Applications”) AND PUBYEAR > 2018 AND PUBYEAR < 2024 AND (LIMIT-TO (DOCTYPE, “ar”)) AND (LIMIT-TO (LANGUAGE, “English”))

The search yielded an initial dataset of 21,141 records. Following the PRISMA framework, records were screened for relevance, with 12,234 records removed due to duplication, irrelevance, or incomplete data. The final dataset comprised 8,907 records included for bibliometric analysis. The extraction process was completed on 7th December 2024.

Bibliometric tools, including VOSviewer and Microsoft Excel, were utilized to analyze the dataset. Key metrics such as citation counts, authorship patterns, institutional contributions, keyword co-occurrence, and geographical distributions were assessed. Visualizations were generated to depict trends and relationships within the dataset, enabling a comprehensive understanding of the research landscape.

This methodology ensures a robust and systematic approach to analyzing the role of AI in revolutionizing healthcare practices and patient outcomes, providing actionable insights for researchers and practitioners.

## 4.0 RESULTS

The results section provides a comprehensive analysis of the research landscape on artificial intelligence (AI) in medicine, emphasizing its evolution, trends, and contributions to healthcare practices and patient outcomes. The findings, derived from a bibliometric analysis of 8,907 records, highlight the significant growth in publications from 2019 to 2023, with a marked increase in 2023. Key metrics such as co-authorship networks, geographical distributions, keyword co-occurrence, and institutional contributions are analyzed to reveal dominant themes, influential contributors, and emerging trends. By exploring these dimensions, this section offers insights into the interdisciplinary and collaborative nature of AI research in medicine, underscoring its critical role in advancing innovative healthcare solutions and addressing complex global challenges.

### 4.1 Increasing number of publications

The results presented highlight the significant growth in the number of publications over the years, reflecting an increasing academic focus on the field. The table provides a year-wise breakdown of documents published, their percentage contributions, and the cumulative percentages, offering insights into publication trends from 2019 to 2023.

**Table 1.** Documents by year of publication trends from 2019 to 2023.

| YEAR | Documents | Percentage (%) | Cumulative Percentage (%) |
| --- | --- | --- | --- |
| 2023 | 2773 | 31.13 | 100.00 |
| 2022 | 2264 | 25.42 | 68.87 |
| 2021 | 1882 | 21.13 | 43.45 |
| 2020 | 1251 | 14.05 | 22.32 |
| 2019 | 737 | 8.27 | 8.27 |

In 2023, the highest number of documents was published, totalling 2,773 and accounting for 31.13% of all publications. This marks a significant surge in academic output, likely driven by advancements in technology and growing research interest. The cumulative percentage for 2023 reached 100%, showcasing the continuous buildup of research over the analyzed period. The year 2022 also saw substantial contributions with 2,264 publications, representing 25.42% of the total and bringing the cumulative percentage to 68.87%. The steady increase continued from 2021 (21.13%, 1,882 documents) and 2020 (14.05%, 1,251 documents), demonstrating consistent growth. The trend began in 2019, with 737 documents accounting for 8.27% of the total.

The cumulative percentage values indicate that by 2022, nearly 70% of all publications had already been produced, pointing to the rapid acceleration of research within a short span. This progression reflects the dynamic nature of the field, with each subsequent year building upon the momentum of the previous one, fostering a vibrant research ecosystem.

### 4.2 Institution with most contribution

The analysis of institutions with the most contributions highlights key players in the research domain, reflecting their active roles in driving scientific advancements. Harvard Medical School stands out as the leading institution, contributing 190 documents, which accounts for 16.46% of the total output. This underscores its pivotal role in fostering cutting-edge research, particularly in areas intersecting medicine and technology.

**Table 2.** Documents by affiliation with the most contributions highlights key players in the research domain.

| AFFILIATION | Documents | Percentage (%) |
| --- | --- | --- |
| Harvard Medical School | 190 | 16.46 |
| University of Toronto | 116 | 10.05 |
| Massachusetts General Hospital | 115 | 9.97 |
| Ministry of Education of the People's Republic of China | 111 | 9.62 |
| King Saud University | 111 | 9.62 |
| Stanford University | 109 | 9.45 |
| Chinese Academy of Sciences | 107 | 9.27 |
| Stanford University School of Medicine | 107 | 9.27 |
| Imperial College London | 94 | 8.15 |
| Brigham and Women's Hospital | 94 | 8.15 |

The University of Toronto ranks second with 116 documents (10.05%), showcasing its significant presence in the global research landscape. Massachusetts General Hospital follows closely with 115 documents (9.97%), reflecting its strong emphasis on clinical research and innovation.

Other prominent contributors include the Ministry of Education of the People’s Republic of China and King Saud University, both contributing 111 documents (9.62%). Their involvement highlights the increasing global diversity in research leadership, with substantial contributions from Asia and the Middle East.

Stanford University (109 documents, 9.45%), the Chinese Academy of Sciences (107 documents, 9.27%), and the Stanford University School of Medicine (107 documents, 9.27%) further emphasize the importance of interdisciplinary collaborations between academic and research institutions. Finally, Imperial College London and Brigham and Women’s Hospital each contributed 94 documents (8.15%), demonstrating their strong commitment to advancing research in healthcare and beyond.

These institutions collectively represent the global of innovation, fostering collaboration across diverse fields to address critical challenges and drive impactful research outcomes. Their contributions highlight the importance of institutional support and resources in achieving sustained research excellence.

### 4.3 Co-occurrence analysis

The analysis of the keyword co-occurrence network highlights the key focus areas and dominant research themes in the domain of artificial intelligence (AI) and healthcare. The most frequently occurring keywords, such as “human” (3,585 occurrences), “article” (3,457 occurrences), and “machine learning” (2,813 occurrences), underscore the centrality of AI technologies and their applications to human-centric medical research and clinical practices.

**Figure 2.**
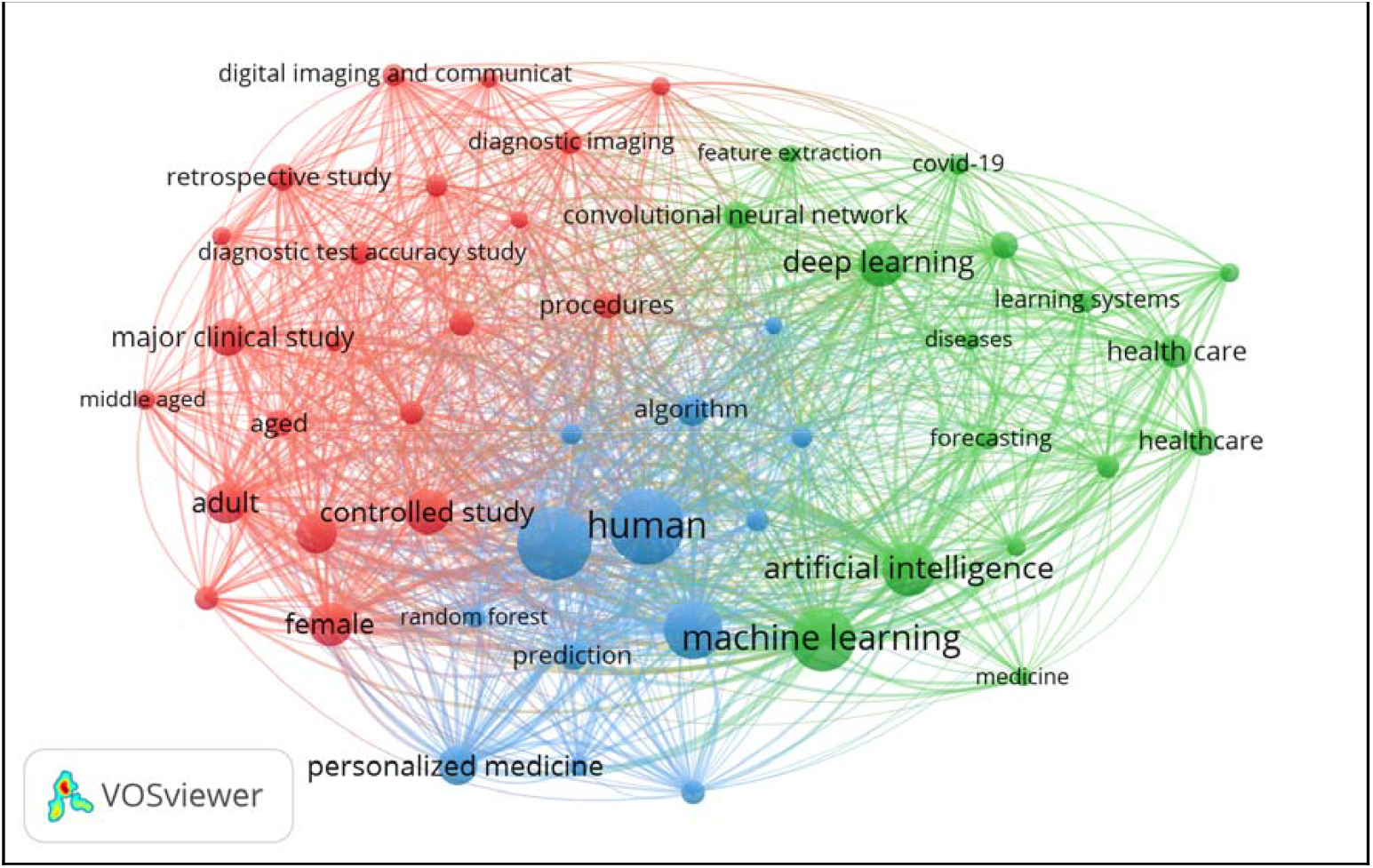
Co-occurrence network of key focus areas and dominant research themes

The network visualization illustrates the interconnectedness between keywords, forming clusters that represent thematic areas. For instance, terms like “deep learning,” “controlled study,” and “personalized medicine” indicate advancements in AI-driven methodologies for tailored healthcare solutions. Similarly, “algorithm,” “health care,” and “major clinical study” reflect the growing emphasis on integrating AI into clinical trials and healthcare systems for improved efficiency and outcomes.

The clustering also highlights emerging areas such as “digital imaging,” “COVID-19,” and “convolutional neural network,” indicating how AI has been rapidly adopted to address contemporary challenges in medicine. Keywords like “forecasting” and “learning systems” point to ongoing research in predictive modeling and intelligent systems for healthcare optimization.

Overall, the analysis underscores the interdisciplinary nature of AI in medicine, bridging areas like diagnostics, imaging, and personalized treatment strategies. The high co-occurrence strength of these keywords reflects a collaborative and integrative approach, driving innovation and addressing critical challenges in modern healthcare. This network provides a roadmap for future research directions, emphasizing the importance of AI in transforming patient care and clinical outcomes.

### 4.4 Co-authorship analysis

The bibliometric data and visualizations provide valuable insights into global research collaboration and its impact in specific academic fields. The co-authorship network, illustrated through VOSviewer, reveals that the United States, China, and India are leading contributors, forming the backbone of global scientific collaboration. Their prominence in the network stems from their significant number of publications and citations, underscoring their influence and active engagement in international partnerships.

**Figure 3.**
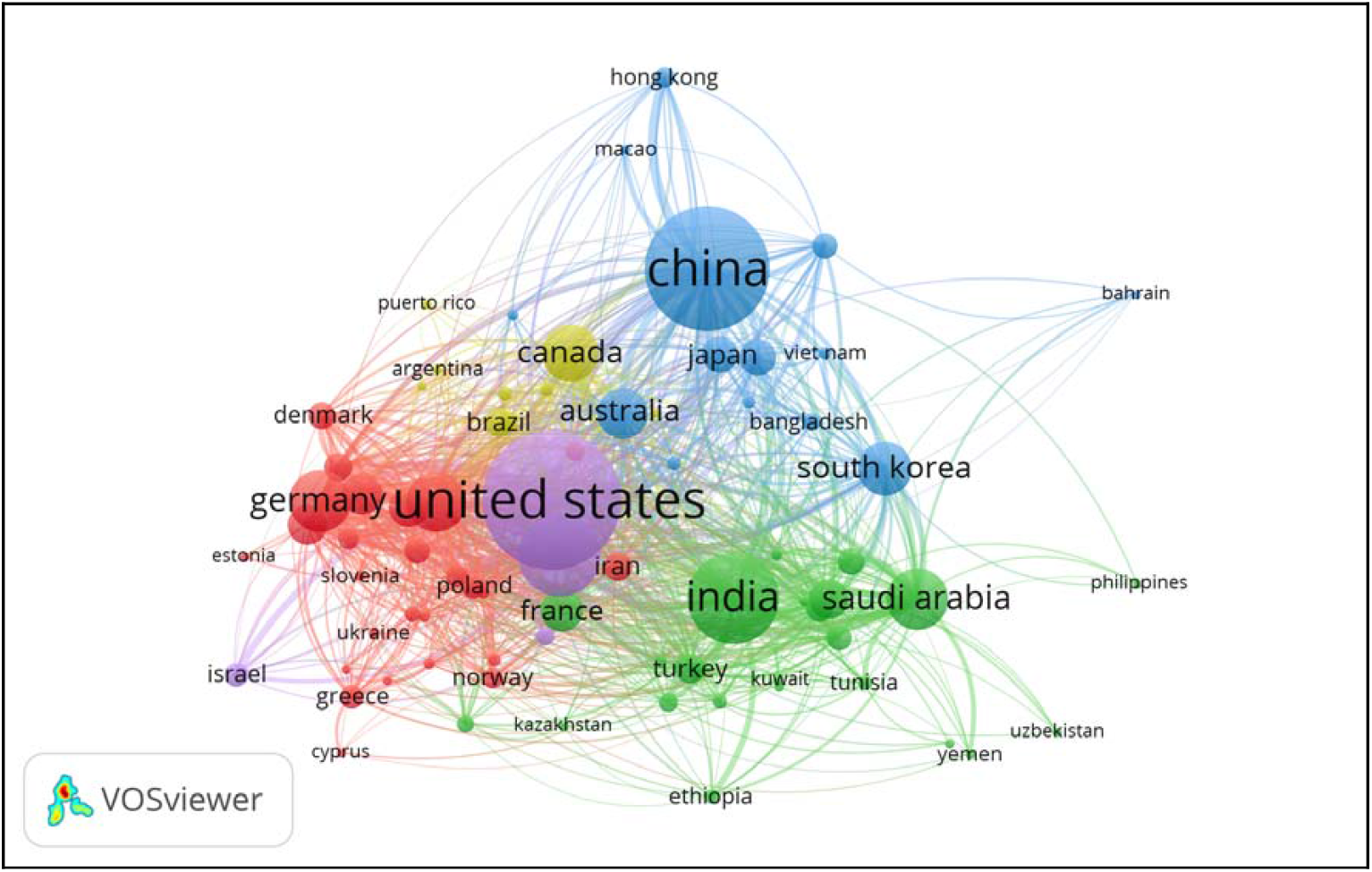
The bibliometric data and visualizations of global research collaboration

The cluster analysis highlights regional and thematic groupings, with countries like Germany, the United Kingdom, and Saudi Arabia forming secondary hubs. These nations act as bridges within their respective clusters, facilitating cross-regional collaboration. The total link strength metric quantifies the depth of these partnerships, emphasizing the interconnectedness of global research. For instance, China’s strong ties with other Asian countries and its increasing collaborations with Western nations reflect its growing role in shaping the research landscape.

Citations and document count further enhance the understanding of impact. The United States leads in both metrics, with 1,475 documents and 22,837 citations, followed by China and India, with substantial contributions of 1,218 documents and 16,170 citations and 716 documents with 12,644 citations, respectively. These figures highlight the quality and reach of their research outputs. Emerging players such as Saudi Arabia and South Korea show significant growth, demonstrating the increasing diversification of global research influence.

Countries like Italy, Canada, and Australia also contribute meaningfully, albeit with lower total outputs compared to the dominant players. Their high citation-to-document ratios indicate a focus on high-quality, impactful research. This distribution reflects the importance of both quantity and quality in global academic influence.

Overall, this analysis demonstrates the dynamic and interconnected nature of global research collaboration. It highlights how leading nations like the United States and China drive innovation while fostering partnerships that enable emerging contributors to play an increasingly prominent role. These findings emphasize the critical role of international cooperation in advancing global research and development.

### 4.5 Highly cited articles

The table of highly cited articles provides a clear view of impactful research in the fields of artificial intelligence, medicine, and healthcare. It highlights notable contributions by leading authors and underscores the growing influence of specific topics in contemporary research. The article *“The Future of Digital Health with Federated Learning”* by N. Rieke et al., published in 2020, leads with 1,219 citations and an average of 304.75 citations per year. This indicates a strong and sustained interest in the application of federated learning to digital health, reflecting its transformative potential in data privacy and AI-driven healthcare solutions.

**Table 3.** Highly cited articles of impactful research in the fields.

| No. | Authors | Title | Year | Cites | CitesPerYear |
| --- | --- | --- | --- | --- | --- |
| 1 | N. Rieke et al. | The future of digital health with federated learning | 2020 | 1219 | 304.75 |
| 2 | D.A. Vyas et al. | Hidden in Plain Sight-Reconsidering the Use of Race Correction in Clinical Algorithms | 2020 | 1109 | 277.25 |
| 3 | J.C. Denny et al. | The "all of us" research program | 2019 | 1002 | 200.4 |
| 4 | E. Tjoa et al. | A Survey on Explainable Artificial Intelligence (XAI): Toward Medical XAI | 2021 | 935 | 311.67 |
| 5 | K. Bera et al. | Artificial intelligence in digital pathology — new tools for diagnosis and precision oncology | 2019 | 891 | 178.2 |
| 6 | M.E. Mayerhoefer et al. | Introduction to radiomics | 2020 | 887 | 221.75 |
| 7 | T. Kobayashi et al. | Ulcerative colitis | 2020 | 881 | 220.25 |
| 8 | C. Longoni et al. | Resistance to Medical Artificial Intelligence | 2019 | 762 | 152.4 |
| 9 | P. Lee et al. | Benefits, Limits, and Risks of GPT-4 as an AI Chatbot for Medicine. | 2023 | 678 | 678 |
| 10 | G.A. Kaissis et al. | Secure, privacy-preserving and federated machine learning in medical imaging | 2020 | 647 | 161.75 |

Similarly, *“Hidden in Plain Sight: Reconsidering the Use of Race Correction in Clinical Algorithms”* by D.A. Vyas et al. (2020), with 1,109 citations and 277.25 citations per year, emphasizes the critical discourse surrounding equity and fairness in medical AI applications. This work’s prominence highlights the ethical considerations integral to AI’s adoption in healthcare.

Other standout articles include *“A Survey on Explainable Artificial Intelligence (XAI): Toward Medical XAI”* by E. Tjoa et al. (2021), which garnered 935 citations with an impressive 311.67 citations per year, showcasing the importance of transparency in AI models for medical decision-making. Articles like *“Introduction to Radiomics”* by M.E. Mayerhoefer et al. (2020) and *“Artificial Intelligence in Digital Pathology”* by K. Bera et al. (2019) reflect the intersection of AI with specialized fields like radiology and oncology.

Lastly, P. Lee et al.’s recent 2023 publication *“Benefits, Limits, and Risks of GPT-4 as an AI Chatbot for Medicine”* achieved remarkable traction, amassing 678 citations within its publication year, showcasing the rapid adoption and critical evaluation of generative AI in healthcare.

This analysis illustrates the diversity of research areas within AI and healthcare, ranging from ethical considerations and explainability to specialized applications in diagnostics and federated learning. It demonstrates how impactful research not only advances knowledge but also shapes the broader discourse on technology and society.

## Conclusion

The analysis of publications across various metrics reveals the dynamic and interdisciplinary nature of research in the domain of artificial intelligence (AI) and its applications in healthcare, technology, and beyond. The increasing number of publications over the years highlights the growing interest and investment in this field, with 2023 marking a significant peak in contributions. The data showcases the pivotal role of leading countries such as the United States, China, and India, alongside major institutions like Harvard Medical School and the University of Toronto, in driving global research efforts.

The co-authorship and keyword networks underline the collaborative and thematic focus of the research, with key areas such as machine learning, deep learning, and personalized medicine shaping the direction of innovation. Journals like *IEEE Access* and the *IEEE Journal of Biomedical and Health Informatics* emerge as prominent platforms for disseminating impactful research. The contributions from top authors such as Rehman, A., and Saba, T., further highlight the importance of individual and institutional efforts in advancing the field.

The diverse subject areas, ranging from medicine and computer science to biochemistry and social sciences, emphasize the interdisciplinary nature of this research. This integrative approach fosters collaboration across fields, driving transformative applications and solutions to complex problems. Overall, the findings illustrate a thriving research ecosystem that continues to expand its scope and impact, paving the way for innovative technologies and methods to address global challenges effectively.

## 5.0 DISCUSSION

The findings of this study reveal significant insights into the role of artificial intelligence (AI) in revolutionizing healthcare practices and improving patient outcomes. The bibliometric analysis highlights a steady increase in publications from 2019 to 2023, reflecting growing academic and industry interest in leveraging AI for medical advancements. This growth aligns with the global push for digital transformation in healthcare, driven by the need for more efficient, accurate, and personalized medical solutions.

### 5.1 Key Trends in AI Applications

AI has demonstrated transformative potential in key areas such as radiomics, genomics, and predictive analytics. Radiomics has enabled more precise and personalized treatment planning, while genomic applications have accelerated the identification of disease markers, paving the way for advancements in precision medicine. The adoption of federated learning frameworks addresses data privacy concerns, allowing for collaborative research without compromising sensitive patient information. These trends indicate a shift toward more integrated, interdisciplinary approaches, combining computational power with domain expertise.

### 5.2 Addressing Challenges

Despite these advancements, several challenges persist in implementing AI in healthcare. Data privacy and security remain significant barriers, as highlighted in the literature. Ensuring confidentiality while enabling seamless data sharing is a critical hurdle that necessitates robust encryption and privacy-preserving technologies. Additionally, algorithmic bias and a lack of transparency raise concerns about fairness and trust in AI systems. The findings underscore the importance of developing explainable AI models that are equitable and reproducible.

Another key challenge is the integration of AI into clinical workflows. Many healthcare institutions lack the infrastructure and expertise to effectively deploy AI technologies. Training healthcare professionals to adapt to these systems is vital for achieving widespread adoption. Addressing these challenges requires collaborative efforts from technologists, clinicians, and policymakers.

### 5.3 Future Directions

The study identifies several avenues for future research and innovation. Advancements in explainable AI and the integration of wearable technologies are expected to enhance patient engagement and real-time monitoring. The findings also emphasize the importance of ethical considerations, particularly in ensuring algorithmic fairness and equitable access to AI-driven healthcare. Standardization of data protocols and interdisciplinary collaboration will be critical in overcoming existing barriers and fostering trust in AI applications.

### 5.4 Implications for Practice

The transformative impact of AI in medicine extends beyond academia, with practical implications for clinical care, operational efficiency, and patient outcomes. By leveraging AI-driven tools, healthcare providers can optimize resource allocation, improve diagnostic accuracy, and deliver more personalized treatments. Policymakers and stakeholders must work together to create a supportive ecosystem that facilitates the safe and effective deployment of AI technologies in healthcare settings.

In conclusion, the findings underscore the immense potential of AI to transform healthcare while highlighting the need to address implementation challenges. By fostering collaboration, prioritizing ethical considerations, and investing in infrastructure and training, the healthcare industry can unlock the full potential of AI to improve global health outcomes.

## 6.0 CONCLUSION

This study demonstrates the profound impact of artificial intelligence (AI) on medicine, highlighting its transformative role in advancing healthcare practices and improving patient outcomes. The findings reveal a significant growth in research output from 2019 to 2023, with key focus areas including radiomics, genomics, and predictive analytics. Innovations such as federated learning and wearable technologies address critical challenges, enabling collaborative research and real-time patient monitoring.

Despite these advancements, challenges persist in ensuring data privacy, overcoming algorithmic bias, and integrating AI into clinical workflows. Addressing these barriers requires ethical considerations, the standardization of data protocols, and interdisciplinary collaboration among technologists, clinicians, and policymakers.

Future research should prioritize the development of explainable and equitable AI systems, fostering trust and ensuring broad access to AI-driven healthcare solutions. By addressing these challenges and leveraging AI’s potential, the healthcare industry can achieve transformative advancements, ultimately improving global health outcomes and enhancing the quality of patient care.

## Data Availability

All data produced are available online at https://doi.org/10.17632/wfdp6t7v2f.1

## Data Availability Statement

The dataset used in this study is publicly available at https://www.kaggle.com/datasets/yudi299/artificial-intelligence-in-medicine-revolutionizi

## Dataset reference

Budi Susilo, Yudi Kurniawan (2025), “Artificial Intelligence in Medicine: Revolutionizing Healthcare Practices and Patient Outcomes”, Mendeley Data, V1, doi: 10.17632/wfdp6t7v2f.1

